# The role of social support in achieving weight loss in adults in the Caribbean aiming to achieve remission of type 2 diabetes: A cross-case analysis

**DOI:** 10.1101/2021.09.13.21263293

**Authors:** Latoya Bartholomew, Nigel Unwin, Cornelia Guell, Karen Bynoe, Madhuvanti M. Murphy

## Abstract

**Background:** Remission of type 2 diabetes through weight loss is possible in a high proportion of persons with a recent diagnosis, but a major challenge is achieving sufficient weight loss.

**Objectives:** In the first study of this type in the Caribbean, we investigated factors associated with successful weight loss in adults in a diabetes remission intervention. We hypothesized that differences in social support may have influenced differences in weight loss achieved by participants in the Barbados Diabetes Reversal Study (BDRS).

**Methods:** A comparative case study was conducted. Quantitative data for the primary outcome measure of weight reduction (the participants’ baseline and 8-month weights) were assessed to identify the 6 participants with the highest and 6 participants with the lowest weight loss. The 8-week (low-calorie diet phase) and 8-month (weight maintenance phase) interview transcripts for each participant were then analysed via qualitative thematic analysis to explore factors related to social support.

**Results:** Informal and formal support were identified for both categories of participants. Cases were similar with respect to their sources of support however dissimilarities were found in (1) the depth of support received; (2) access to supportive environments and (3) diversity of social supportive networks. Participants in the top weight loss group reported consistency in the levels of support received over the low-calorie diet and weight maintenance phases of the study while the converse was true for those of the bottom weight loss group.

**Conclusion:** Study findings suggest that individuals aiming at type 2 diabetes remission benefit from strong social support networks. These networks provide tangible assistance and facilitate the sharing and discussion of strategies for weight reduction. Future studies should facilitate in-depth understanding of how formal and informal supportive networks can aid sustained dietary diabetes remission and long-term weight maintenance.

## Introduction

Diabetes is a chronic long-term condition and one of the fastest growing health concerns of the twenty-first century. In 2019, it was estimated that approximately four hundred and sixty-three million adults 20-79 years of age were living with the condition worldwide; a threefold increase from the one hundred and fifty-one million global estimate in 2000 (1). The increase in the prevalence of diabetes is largely due to the rise in type 2 diabetes which accounts for approximately ninety percent of global cases (2). The burdens of the disease are particularly severe for low- and middle-income countries where approximately seventy-seven percent of all individuals with diabetes reside (3). One of the fastest growing increases in the prevalence of type 2 diabetes has occurred in the region of the Americas and Caribbean with the most dramatic rise taking place in the countries of the English-speaking Caribbean (4). In Barbados, the Health of the Nation (HoTN) epidemiological survey, found that approximately nineteen percent of adults have diabetes with the prevalence for those sixty-five years and older being as much as forty-six percent (5).

Although diabetes is regarded as a progressive lifelong disorder, research now suggests that its progress can be halted, and remission achieved; the findings of the Diabetes Remission Clinical Trial (DiRECT) in the UK provide compelling evidence of this. In particular, the study suggests that individuals living with type 2 diabetes for less than six years can return to long-term non-diabetic glucose control, off all glucose lowering medication, through substantial and maintained weight loss (6). Over a twelve-month period of sustained weight loss, eighty-six percent of participants with at least fifteen kilograms weight loss and seventy-three percent with ten kilograms weight loss or more achieved remission (7). The researchers’ results were further substantiated as their two-year findings suggest that continuous remission is possible as seventy percent of participants who maintained weight loss of more than fifteen kilograms continued in a non-diabetic state over a twenty-four month period (8).

As suggested by the data of the DiRECT study, achieving, and maintaining weight loss is a critical factor for the remission of type 2 diabetes. The rapid weight loss achieved by the uptake of a very low-calorie diet (VLCD) such as that used in the DiRECT study and the associated health improvements one experiences serve as a major motive for diet adherence(9-11). However, one can encounter several threats (such as tempting situations and environments, emotional distress, and high costs of meal replacements) which can negatively impact one’s ability to adhere to such a diet. It is thus critical that behaviour change strategies are deployed to increase adherence to the VLCD. The implementation of behavioural strategies such as the removal of temptations from immediate environments, avoidance of tempting social situations and places, and meal planning are critical for countering barriers to VLCD (9, 10). Furthermore, though individual behavioural factors may fundamentally impact one’s health outcomes, interpersonal and organizational factors can also ultimately affect one’s health behaviours (12-14). One such factor is a supportive social network which facilitates adherence to VLCD (9-11) and has been increasingly recognized as having an integral role in enhancing health outcomes for persons with a wide range of conditions (15-20).

Social support is a complex concept which has been a subject of examination over multiple disciplines. For the purpose of this study we used the following definition: “a transactional communicative process, including verbal and or non-verbal communication that aims to improve an individual’s feelings of coping, competence, belonging and or esteem”(21). Through this interpersonal transaction, persons are afforded (i) emotional support which addresses an individual’s emotional or affective needs; (ii) esteem support which focuses on communication that encourages an individual to take needed actions despite perceived difficulties; (iii) information support which examines communication that provides useful and necessary information; (iv) tangible support which assesses the physical assistance offered to those in need and (v) network support which alludes to the importance of an individual belonging to a network where persons are available to provide all or any of the abovementioned types of support. An examination of the utility of social support in the remission of type two diabetes is thus timely as it extends beyond an individual behavioural focus and thus illuminates the role of the ecological and social context of self-management support.

Although one’s social network can play a pivotal role in maintaining one’s well-being, the structure of the network must match the structure of a task that provides support, if there is to be a positive correlation between social support and health (15). Research has demonstrated that support provided by primary groups inclusive of families and friends has had a positive impact on diabetes self-care and management such as physical activity and weight loss (22-24), while secondary or constructed group support resulted in improvements in HbA1C levels (22). Whilst these studies point to the positive influence of a supportive network, it can be argued that persons who have more diversified supportive networks have greater access to health support and thus have a greater propensity to experience positive health outcomes (17, 25, 26).

This study therefore explored participants’ access to supportive resources and variations in the support received, in Barbados, with the aim of understanding its role in the reduction of the primary outcome measure of weight.

## Methodology

This analysis formed part of the Barbados Diabetes Reversal Study (BDRS); details of the methods have been previously published (27). The BDRS utilized a mix methods framework to examine the acceptability and transferability of a very low-calorie diet and structured long-term support for reversing type 2 diabetes in Barbados. This feasibility study spanned an eight-month period with the first two months being for the very low-calorie diet phase, followed by a six-month structured weight maintenance phase. Twenty-five Barbadians who were diagnosed with type 2 diabetes for six years or less between the ages of 20-69 with a body mass index of ≥ 27 kg/m^2^ participated in the study. Ethical approval was obtained from the University of the West Indies Institutional Review Board and the Ministry of Health of the Government of Barbados. Written informed consent was secured from all participants.

Using a ‘before and after’ study design and standard meal tests, the primary outcome measures of a reduction in weight, fasting glucose, insulin resistance and an increase in insulin secretion were assessed. Participants stopped their use of glucose-lowering medication on day one of the very low-calorie diet. The researchers also took note of secondary outcome measures such as reductions in the participants’ waist circumference, blood pressure, and adverse lipid profile levels. Semi-structured interviews were also conducted during the three main data collection points (baseline, low-calorie diet phase/ eight-week, end of study/ eight-month) to identify and understand the challenges and facilitators participants experienced complying with the intervention.

### Setting

Barbados is the most easterly Caribbean island covering an area of 431 km^2^ (28). Healthcare on the island is characterised as a two-tiered system with a publicly funded health system operating in conjunction with numerous private clinics (29). The Ministry of Health delivers and regulates the three levels of healthcare (primary, secondary, and tertiary) in the public sector. Primary care for diabetes is delivered through a network of nine Ministry of Health polyclinics which are easily accessible in their respective catchment areas or by private general practitioners for a fee (30, 31). Those patients who have either uncontrolled diabetes or are at risk of complications are referred from the primary healthcare system to the Barbados Diabetes Foundation’s speciality clinic (30). Patients are managed at a fee for a six-month period after which time they are once again referred to the primary healthcare system. In addition, the Queen Elizabeth Hospital is the main provider of acute secondary and tertiary care (32). Through the Barbados Drug Benefit Service, patients accessing care in the public and private sectors can secure at any one time a maximum of one-month’s supply of free medication (31).

Despite the Government’s effort to ensure healthcare is accessible to all citizens there are still challenges it needs to overcome. Some of these challenges include (i) lack of confidence in the primary care services that limits access to care; (ii) long waiting times to be seen in the polyclinics; long waiting lists at the Queen Elizabeth Hospital for complex medical procedures and (iv) medication supply problems in the polyclinics (31, 32), all which affect the level of support received by patients.

### Study Design

A comparative case study design was chosen as it offered the researchers the opportunity to explore and synthesize similarities and differences embedded in multiple cases. The study was guided by the hypothesis that: “BDRS participants who reported having consistently more social support over the course of the study experienced greater reductions in their weight.” Therefore, the researchers sought to discover and garner rich descriptive data on; (i) the types of social support reported by the participants; and (ii) variations in social support received over the course of the study. Resultantly, three questions were posed to further direct the scope of this paper. They are as follows

1. How do the BDRS participants who experienced the greatest weight loss describe social support received during the course of the intervention compared to participants who lost the least weight?
2. What similarities and differences in social support were reported by the participants?
3. Was there variation in social support received by the participants over the duration (i.e. low-calorie diet and weight maintenance phases) of the intervention?

### Sampling

Purposive sampling was applied to identify the cases to be examined. Quantitative data collected for the twenty-four participants who completed the study were reviewed to identify those who experienced improvements as well as those with little to no improvements in their weight. The participants’ baseline and eight-month weights were evaluated and the top six (high weight loss or HWL) and bottom six (low weight loss or LWL) participants were then chosen (n=12). This choice was based on the overall percentage change of weight loss calculated for all participants. Table 1 provides an overview of participants’ weight at the baseline and end of the study.

**Table 1.** Percentage change in weight for top 6 and bottom 6 participants between baseline and eight-month measures

| ID # | Baseline weight | 8-month weight | % change |
| --- | --- | --- | --- |
| High Weight Loss (HWL)- (kg) |  |  |  |
| HWL 1 | 121.5 | 102.3 | 15.80 |
| HWL 2 | 94.3 | 81 | 14.10 |
| HWL 3 | 106.8 | 92.1 | 13.76 |
| HWL 4 | 75.8 | 65.4 | 13.72 |
| HWL 5 | 111.2 | 96 | 13.66 |
| HWL 6 | 91.4 | 79.1 | 13.45 |
| Low Weight Loss (LWL)- (kg) |  |  |  |
| LWL 1 | 92 | 90.9 | 1.19 |
| LWL 2 | 73.6 | 72.3 | 1.76 |
| LWL 3 | 86 | 84.2 | 2.09 |
| LWL 4 | 78 | 76.3 | 2.17 |
| LWL 5 | 139 | 135.7 | 2.37 |
| LWL 6 | 79.8 | 82.6 | *3.5 + |

(+) increase in outcome measurement

Multiple cases were chosen as the researchers attempted to gather data on social support that was representative of the experiences of those respondents who achieved the most and least weight loss. As espoused by Yin (33), multiple cases ultimately provide convincing support of the proposition guiding the study if similar results are collectively garnered. It must be noted however the researchers are by no means attempting to propose statistical generalizations but rather analytical generalizations which elucidate the experiences of the BDRS participants.

### Data collection and analysis

For this paper specific attention was given to the eight-week (low-calorie diet phase) and eight-month (end of study) BDRS interviews. This choice was made as these interviews examined the role of social support at critical junctures for the participants. More specifically, the interview guides focused on topics such as (i) barriers and facilitators experienced during the study; (ii) types of social support received, (iii) sources of social support and (iv) variation in social support received. Interviews were conducted by two members of the research team L.B and K.B.

All interviews, apart from two, were audio-taped using a digital recorder and transcribed verbatim using the Express Scribe software. In the instance where the participant preferred not to be recorded detailed notes were taken. All information highlighting the identities of the participants was removed and pseudonyms assigned.

The eight-week and eight-month transcripts of the HWL and LWL participants were inductively examined through thematic analysis. Such analysis enables one to identify, code and categorize common themes within the data (34). A total of twenty-four transcripts were independently coded using the ATLAS.ti 8.0 data analysis software (ATLAS.ti Scientific Software Development GmbH, Berlin Germany). This was then followed by a cross-case analysis to discern the similarities and differences in themes between cases. Verbatim quotes which epitomized the emergent themes are highlighted in the subsequent section.

## Results

A total of twelve participants were included in this study. Tables 2 and 3 provide an overview of their demographic and outcome measurements. Four participants experienced concomitant reductions in three outcome measures; seven experienced reductions in two outcome measures but increases in another; 1 participant had an inconclusive eight-month HbA1c measurement but reductions in the other outcome measures.

**Table 2.** Comparison of baseline characteristics and weight loss in the high weight loss and low weight loss participants

| **Demographic data** |  |  |
| --- | --- | --- |
| Average age of the 12 participants: |  | 46.1 (12.9) |
| Sex: | 7(Females) | 5(Males) |
| Years living with diabetes: |  | (2.6) |
|  | **6 HWL** | **6 LWL** |
| Average age: | 50.2 (7.78) | 42 (16.26) |
| Sex: | 4 (Females) | 3 (Females) |
|  | 2 (Males) | 3 (Males) |
| **Weight (kg)** |  |  |
| Baseline | 100.16 (16.28) | 91.4 (24.19) |
| 8-month | 85.98 (13.41) | 90.33 (23.14) |

N= 12. Results are presented as mean ± SD.

**Table 3.** Mean fasting glucose and HbA1c from baseline to end of study for the high weight loss and low weight loss participants

|  | **6 HWL** | **6 LWL** |
| --- | --- | --- |
| **Fasting glucose (mmol/l)** |  |  |
| Baseline | 8.01 (1.26) | 9.4 (1.71) |
| 8-month | 6.98 (0.63) | 8.03 (0.77) |
| **HbA1c (%)** |  |  |
| Baseline | 7.75 (1.64) | 7.91 (1.63) |
| 8-month | 7.22 (0.54) | 7.8 (1.43) |

N= 12. Results are presented as mean ± SD.

### Sources of support

Participants reported a range of sources of social support. There was consensus among participants that they relied heavily on members of their primary and secondary groups for social support during the intervention. The data suggests that informal support is classified as any assistance provided by members of the participants’ primary and secondary groups. These groups consisted of their family members, close friends, co-workers, and other participants. While family members were highlighted as being a major source of support for HWL participants three LWL participants reported lack of support related to perceptions of long-term success of the intervention. In the instance of the latter, participants’ family members expressed scepticism of their ability to successfully complete the intervention and were therefore deemed to be a source of “negative support”. They also noted that their family members failed to provide them with encouragement, and even ridiculed their newly adopted eating habits. One participant recounted:

> “… everybody saying, ‘man you go soon dead eating like a bird’. **[Male participant, low weight loss group, 8-week interview]**

He goes on to further state:

> “…I alone had to struggle for myself because umm talking to others about diabetic stuff, some of them don’t understand the reasoning… [I] have to be strong because people in the house that I live with don’t really understand. Everybody feels that when you got diabetes in the next couple of months you going dead…But people don’t really help. They need to get more involved than this.” **[Male participant, low weight loss group, 8-month interview]**

Participants who lacked social support from their family either depended on their close friends, significant other and or themselves as they dealt with the challenges of being on the low-calorie diet.

Interestingly, the participants created their own informal support network through the creation of a WhatsApp chat group with the sole purpose of motivating one another especially during the weight maintenance phase of the study and points when they experienced an upward shift in their measurements. Both HWL and LWL participants noted that their involvement in this chat group gave them a sense of accountability to one another. It also gave them a level of reassurance that they were not the only one having trouble engaging in and maintaining the recommended behaviour.

There was also consensus among all participants of the importance of formal support networks (members of the BDRS research team and their private doctors) with the BDRS team being a key source of formal support. Participants reported that the intervention’s structure made it easier for them to manage their illness. They expressed similar opinions on the utility of the weekly visits with the chief clinical researcher and weekly calls from the other members of the team. These weekly check-ins they agreed, gave them a greater sense of accountability especially during the first eight weeks of the study when they were faced with more intense challenges.

All participants expressed their preference for the structured and sustained support received during the first phase of the study. Although the team intentionally reduced the number of visits and calls during the second phase, participants in the low weight loss group recognized it as a change in the level of social support. On the other hand, those in the high weight loss group suggested more formal support could have been given to persons who were having a difficult time sticking to the recommended behaviour.

In addition to the medical support and advice given by the team, the participants referenced the formal support gained from their private physicians. Social support in this instance took the form of encouragement, check-ups, and provision of further information on ways to effectively manage their diabetes. There was mixed feedback from participants when asked about their relationship with their personal doctors. Those in the low weight loss group expressed that their relationship with their private doctor was non-existent and they did not get the type of support they thought was needed from them. One participant stated:

> “I attend the polyclinic and it is somewhat a rush that you could only get in a small amount of questions with the doctor…But here at the clinic with [the BDRS chief clinical investigator] you get more one to one and in-depth analysis- ‘well do this, this is what the purpose about, this is what you should be looking for and aiming for’. So [the BDRS chief clinical investigator] always motivates the person. So, I mean doctors, I really don’t care much for doctors.” **[Male participant, low weight loss group, 8-week interview]**

Nonetheless, it was found that those persons who had a more diversified social support network (a mix of formal and informal support) tended to experience greater positive results throughout the study. This was especially the case for the participant topping the weight loss category as he expressed his gratitude for having the assistance of his family members, friends, other participants, members of the BDRS team and private doctor.

### Choice of support

Three recurrent sub-themes were identified with respect to factors that influenced the participants’ choice of persons to whom they relied on for social support during the study. Privacy was one factor consistently reported among all participants. As demonstrated in the quote below, they referenced the fact that their illness was a private matter and they felt more comfortable sharing their experiences with and relying on their immediate family and close friends for support.

> “Umm you could say because you’re going through this whole programme is a kind of, how do I say, ‘a private thing’, so you would feel more comfortable with persons that are closer to you; your friends, close friends and family. So then you tend to lean on those persons more so than other people; not that other persons wouldn’t know, but you would tend more to lean on persons closer to you for support because I think it’s a private personal thing that you’re going through and battling with. **[Female participant, low weight loss group, 8-month interview]**

The data also suggest that the participants relied heavily on persons who consistently provided them with support prior to the commencement of the intervention. Again, they referenced the support obtained from close family and friends, alluding to a “trusted circle” within their informal support network. Participants expressed reservations about sharing any information about the intervention with persons who fell outside of their immediate circle of close family and friends. The strong level of trust and security fostered within this circle made it easier for participants to disclose information about their experiences in the study.

Lastly, expertise arose as an important factor influencing their choice of social support. Some reported that choice of social support was based on the level of knowledge and expertise of their social support provider(s). As such, these participants turned to formal support networks such as their private medical practitioner and members of the research team for advice and guidance on the best steps to take to overcome their challenges and maintain their positive behaviour change.

### Depth in support

The depth of support received over the course of the study was a major difference observed between both groups and went beyond concepts such as words of encouragement. Such variation was found particularly in the consistency of dietetic support reported by the participants. There was consensus amongst the HWL participants that their family members played a critical role in purchasing and preparing their recommended meals. Furthermore, some family members and close friends attempted a variation of their diet and even joined them in exercise sessions to encourage and motivate them to maintain the positive behaviour change.

It was also reported that the participants’ food consumption and portion sizes were monitored by family members and close friends. This form of dietetic support however was largely reported by participants who struggled with their weight loss. They noted a decline in the dietetic support received over the course of the study. Particularly, LWL participants noted that their supporters provided assistance more enthusiastically at the beginning of the study.

Differences were also noted in the depth of support given by the participants’ co-workers. Those who excelled in their weight loss reported that their co-workers implemented a healthy lifestyle challenge mirroring their positive behaviour change. The participant topping the HWL group recounted:

> “…at work, there are like 7 of us. A lot of them have started eating healthy. Coca Cola was my best friend, the regular coke, and I use to consume, even I knew I use to consume too much sometimes. At work now you will hardly ever see a coke because we talk and started to go for more umm healthy foods and stuff like that and exercise. So right now, except for one of the guys, the other 6 of us exercise on a regular basis.” **[Male participant, high weight loss group, 8-month interview]**

Conversely, for those in the low weight loss group, support took the form of encouragement and monitoring the participants’ eating. This support from co-workers, allowed for continuous support from informal networks, throughout the day and in different settings.

### Unsupportive environments

The participants’ ability to meet the study’s requirements was impacted by expectations they deemed unaccommodating in their physical and wider social environments; for example, dinning out and social occasions with family, friends. and work colleagues. Though this was not the experience of most cases, unsupportive environments and behaviours were more frequently referenced by those in the low weight loss group. Negative comments about their rapid weight loss, food consumption choices and patterns and ability to successfully complete the study were just a few comments participants dealt with over the course of the intervention. Interestingly, the male participants in the aforementioned group appeared to be the recipients of such negative comments especially in the home and social environments.

> “The past 4 months I struggle for myself because talking to others about diabetic stuff, some of them don’t understand the reasoning. Then you got people in the community, certain friends saying, ‘man you look bad, you gone soon dead’. Some people went as far as saying, well the popular HIV thing. They say ‘you got the pog’… Everybody feel that when you got diabetes [it] is something that in the next couple of months you going dead. But people don’t really help.” **[Male participant, low weight loss group, 8-month interview]**

Recreational facilities also proved to be problematic for participants. One participant of the low weight loss group coined the phrase “diabetic discrimination” to express the challenges she encountered at the movie theatre where she was denied entry for taking along her Glucerna shake. Despite her attempts to explain her reasons for having the shake, it was only after she threatened to take action should she fall ill while at the establishment, that management granted her access to the facility.

Moreover, when socializing, all participants were forced to address peer pressure to consume normal foods. The slightest resistance shown was perceived as them severing cultural norms. Although it was anticipated that participants would at one point or another break their diet, the data suggests that persons in the high weight loss group were better equipped to address the peer pressure they experienced while on their restricted diets. In general, they appeared to have a higher level of self-determination and will power to maintain the recommended behaviour.

> “I’ll find other options that will make [my boyfriend] feel comfortable with me being social because he has this thing when we go to people’s houses and I say, ‘thanks no’, [he says] ‘you can’t tell people no, they invite you to their house and offer you something, you have to take it. They feel that you think what they have is not good enough.’ So, at that point I started telling people ‘thanks, no, I’m a diabetic, I can’t take that.’ It’s actually been easier in the last 8 weeks because I could say ‘I am part of the Barbados Diabetes Reversal Study; this is why I cannot accept what you’re offering me. It has made it lot easier than just saying I’m diabetic you know.” **[Female participant, high weight loss group, 8-month]**

Finally, the work environment presented challenges for the maintenance of the participants’ healthy habits. Cases within the low weight loss group commonly reported instances when they skipped meals or purchased unhealthier food on the go as they dealt with the demands of their busy schedules. The 6 HWL participants however, reported their ability to pre plan and prepare their meals, along with the support of their colleagues, made it easier for them to address the challenges associated with the work environment.

## Discussion

This qualitative enquiry employed a comparative case study design to assess the differences and similarities in the types and levels of support received by the participants of the Barbados Diabetes Reversal Study. The data suggest that though all participants noted receiving both formal and informal support from similar sources, those who excelled in losing considerable weight received a greater depth of support, had access to more supportive environments and a varied social support network.

### Findings in context

Having access to a varied network was critical as it was a precursor for the quality and depth of support participants received. This notion is supported by Dykstra (35) who posited that one’s social network composition is a powerful predictor of the quantity, quality and types of support one has access to. It was therefore not surprising that participants who excelled with their weight loss reported sourcing support from a network inclusive of family members, friends, co-workers, private doctors, and other members of the study. Mattson and Hall (21) further argue that network support is critical as an individual’s inclusion in a network affords them the opportunity to gain access to various types or combination of types of support. With access to such heterogeneous networks, the top six participants were therefore constantly provided with different types of assistance throughout the course of the intervention. The converse is true for the low weight loss participants as some had access to more restricted networks where support was obtained from friends, other participants and or co-workers for example.

It was clear that family members were crucial informal support providers for most participants. They were instrumental in providing participants with esteem support in the form of encouragement and compliments and tangible support by purchasing and preparing their meals for example. Pinelli et al (23) argue that support from this natural group is an effective strategy to enhance the adoption of diabetes preventative behaviours. Our data reinforces this argument as it suggests that those individuals whose family members were actively involved and supportive of their behaviour change approaches and strategies were more successful in losing and maintaining their weight. More importantly, it became easier for them to maintain their newly adopted habits as their family members also adjusted their eating and exercise practices in solidarity of their positive behaviour change.

Although one may argue that it is expected that such support is provided by this primary group, this was not the experience for three participants of the low weight loss group. For these individuals, the positive social exchanges expected from one’s family members were not forthcoming. In particular, the negative comments received, and their unsupportive home environment served as a deterrent for the recommended behaviour. Though based on their accounts they appeared to be a bit more equipped to handle derogatory comments, the same cannot be said for their ability to manage temptations present, especially as their family continued to maintain their behaviours without any regard for the impact it had on them. A disinterested or uninvolved family thus increases the likelihood of failure for individuals in their bid for weight loss (36). Nonetheless, as suggested by Verheijden et al (37) bringing new sources of support such as peers into action when social support from natural groups such as the family is insufficient can prove to be helpful. This strategy was predominantly useful for these three participants as they relied on the assistance of their peers and other participants.

It is well acknowledged that both formal and informal networks are powerful tools in encouraging positive health behaviour (25). It was therefore not surprising that participants served as another critical source of informal support for one another. Their need to create a mutual support group was intensified as they approached the end of the eight-week diet where consistent structured support from the members of the research team came to an end. Consequently, they formed a WhatsApp chat group as a place of social acceptance where they encouraged each other as they returned to a sense of normalcy with their food consumption. This mediated group not only served as a forum for helping each other discover new ways of coping and addressing the challenges faced but it also served as a source of accountability. Cases from both categories noted that having access to such a group would serve as further motivation for them to maintain the positive behaviour change post intervention. The utility of this group however diminished as they transitioned through the beginning of the weight maintenance phase to the end of the study. All participants noted that by the end of the study, members’ focus shifted from diabetes and weight management strategies to unrelated topics. Resultantly, it can be argued that the utility of such a support group is hinged on having a person or group of people who are willing to take the responsibility of up keeping the group’s overarching aims and objectives.

Based on the participants’ narratives it was also clear that they had a high sense of accountability towards the BDRS team, more so the chief clinical investigator. This high sense of accountability worked in their favour especially during the eight-week phase of the study where they were consistently monitored as they adjusted to their new lifestyles. Interestingly, the participants who struggled with their weight loss expressed a desire for greater formal support. They attributed the short duration of structured support as a reason for their weight gain. Although the structured support was purposefully reduced with the intention of giving participants full responsibility for the maintenance of their lifestyle changes, they argued that continued formal support from the team was necessary for securing a successful health outcome and by extension maintaining their weight loss. The HWL participants however were less inclined to make note of the duration of the structured support provided by the BDRS team. Like their fellow participants some were of the view that more support could have been given to those persons who experienced greater difficulty adhering to the recommended behaviour change.

Research suggests that that continuous professional support in weight maintenance has often been found to improve treatment outcome (38-40). This professional support is argued to be pivotal in enhancing vigilance and motivation. One can however argue that such prolonged support may work to the detriment of the participant as a dependency syndrome can ultimately develop where their actions are predicated on the fact that someone else takes responsibility for their behaviour change. Alternatively, the aim of any professional or formal support provider should be to support the patient’s full responsibility for their lifestyle changes through the provision of tools, skills and tactics that can be incorporated into their daily regimes (41-46). In this vein therefore, the individual is empowered and gain a level of confidence in their ability to manage their weight and by extension their illness. Furthermore, this necessitates more proactive and involved physician support especially from the polyclinic setting where several cases from the LWL group sourced medical assistance. The healthcare team at such institutions should aim to provide practical support, where agreed on goals are set and action plans are developed. This type of practical support is an integral part of the doctor-patient relationship and is an effective part of physician support for diabetes self-management (42, 47, 48).

Though social support may have an indispensable role in the remission of one’s type 2 diabetes, a high level of self-efficacy is required to successfully manage the illness (9, 25, 41, 49, 50). Our findings are consistent with the aforementioned authors’ argument as participants who did particularly well appeared to be more conscious of their food intake and weight related behaviours. More specifically, the six HWL participants took responsibility for their lifestyle changes by paying greater attention to the nutritional content of foods consumed, monitoring their portion sizes, and incorporating physical activity into their daily routine. Additionally, our findings suggest that those with a higher level of self-efficacy were better equipped to address the challenges present in unsupportive social and work environments. In the instance of latter, the difference between both groups laid in the fact that the HWL participants pre-planned and pre-packaged their meals and snacks which they took with them thereby effectively addressing the conflicts in meeting the demands of their busy work schedules and their behaviour change. With respect to the social environment, the participants all agreed that they experienced a sense of otherness as their explanations for not wanting to partake in food and drinks served at gatherings were met with negativity. In most instances they were ridiculed and accused of breaking the cultural norms that guide social gatherings. Those cases who displayed a high level of determination and who were unafraid to divulge their involvement in the study feared better in addressing the pressures of the social environment.

### New perspectives

The findings of this study make a case for the inclusion of heterogeneous support networks in future Caribbean based interventions aimed at type 2 diabetes remission. Long-term weight loss and management appears to be more attainable once one has access to various combinations of social support from one’s formal and informal networks. More importantly, it is crucial that the members of one’s natural group, particularly one’s family members, are included and given active roles in future interventions. As demonstrated by the data, they provide tangible and esteem support which may increase one’s propensity to adopt the requisite behaviour change for substantial sustained weight loss. The continuous provision of informal support also proved useful as participants navigated and addressed challenges within their work and social environments. Researchers can further examine the utility of involving supportive co-workers and peers in future interventions especially in instances where immediate family members are deemed to be unsupportive by participants. Ideally, future researchers should aim to engage a variety of support providers across different social groups within the participants’ informal networks to ensure there is continuity in social support provided in all settings.

Furthermore, given that the BDRS participants spoke highly of the utility of a mutual support group, it would be wise if a similar approach is adopted in future studies. Such a group may prove more effective if it is managed by members of the research team for the duration of the intervention. Consequently, participants would be provided with the formal support and the sense of accountability deemed necessary for the maintenance of their behaviour change. The aim here, however, must be to empower individuals with the tools and skills needed for the maintenance of the requisite behaviour change long after the completion of the intervention.

### Strengths and limitations

This comparative study provides insight into the types and levels of informal and formal social support received by participants of a type 2 diabetes remission intervention. We identified that access to heterogenous support networks (inclusive of family members, friends, co-workers, research team members and private doctors) and the increasing magnitude of support provided, positively impact dietary adherence and weight loss outcomes. This magnitude of support is contingent upon a combination of support actors and continuity of support throughout the intervention. Our findings are however based on the narratives of 12 participants (HWL and LWL); it is possible that those participants who experienced moderate weight loss who are not included in this analysis may have had different experiences than those highlighted here.

## Conclusion

Social support played an integral role in the weight management intervention for remission of type 2 diabetes within a Barbadian context. Having access to varied networks and supportive environments afford one the opportunity to gain access to different forms and combinations of informal and formal support. Although these prove to be useful, a high level of self-efficacy is critical for the maintenance of the behaviours required to maintain substantial weight loss and by extension reverse one’s type 2 diabetes.

## Data Availability

Data in this study comes from a database maintained by the Brazilian Ministry of Health (MoH), which provided the anonymized database for this analyses.

## Acknowledgments

The Barbados Diabetes Reversal Study was funded by a grant from Virgin Unite. We therefore thank Richard Branson for his support. Professor Roy Taylor was the co-principal investigator for the BDRS, visiting Barbados to help establish the study and in particular the protocols for the very low-calorie diet phase and the metabolic outcome measures. For their professional input, we thank Dr. Selvi Jeyaseelan and the late Dr. Charles Taylor of the Faculty of Medical Science, the University of the West Indies, Cave Hill Campus. We thank Krystal Boyea, Melissa Abed and Andre Greenidge who assisted with data collection. Finally, we wish to acknowledge the contributions of the Barbados Diabetes Association, the Barbados Diabetes Foundation, and the George Alleyne Chronic Disease Research Centre.

